# Disseminating sleep education to graduate psychology programs online: A knowledge translation study to improve the management of insomnia

**DOI:** 10.1101/2023.02.23.23286389

**Authors:** Hailey Meaklim, Lisa J. Meltzer, Imogen C. Rehm, Moira F. Junge, Melissa Monfries, Gerard A. Kennedy, Romola S. Bucks, Marnie Graco, Melinda L. Jackson

**Affiliations:** Turner Institute for Brain and Mental Health, School of Psychological Sciences, Monash University, Clayton, Australia; Institute for Breathing and Sleep, Austin Health, Victoria, Australia; National Jewish Health, Denver, USA; Nyxeos Consulting, Denver, USA; College of Health and Biomedicine, Victoria University, Melbourne, Australia; Sleep Health Foundation, East Melbourne, Australia; School of Health and Biomedical Sciences, RMIT University, Bundoora, Australia; Institute of Health and Wellbeing, Federation University, Ballarat, Australia; Schools of Psychological Science and Population and Global Health, University of Western Australia, Western Australia, Australia; Department of Physiotherapy, University of Melbourne, Vic, Australia

**Keywords:** Insomnia, Sleep education, CBT-I, RE-AIM, Implementation, Knowledge Translation, Mental Health, Psychology training, Psychotherapy, Medical Education

## Abstract

**Study objectives:** Despite the negative impact of poor sleep on mental health, evidence-based insomnia management guidelines have not been translated into routine mental healthcare. Here, we evaluate a state-wide knowledge translation effort to disseminate sleep and insomnia education to graduate psychology programs online using the RE-AIM (Reach, Effectiveness, Adoption, Implementation, and Maintenance) evaluation framework.

**Methods:** Using a non-randomized waitlist control design, graduate psychology students attended a validated six-hour online sleep education workshop delivered as part of their graduate psychology program in Victoria, Australia. Sleep knowledge, attitudes, and practice assessments were conducted pre- and post-program, with long-term feedback collected at 12 months.

**Results:** Seven out of ten graduate psychology programs adopted the workshop (adoption rate = 70%). The workshop reached 313 graduate students, with a research participation rate of 81%. The workshop was effective at improving students’ sleep knowledge and self-efficacy to manage sleep disturbances using Cognitive Behavioral Therapy for Insomnia (CBT-I), compared to the waitlist control with medium-to-large effect sizes (all *p* <.001). Implementation feedback was positive, with 96% of students rating the workshop as very good-to-excellent. Twelve-month maintenance data demonstrated that 83% of students had used the sleep knowledge/skills learned in the workshop in their clinical practice. However, more practical training is required to achieve CBT-I competency.

**Conclusion:** Online sleep education workshops can be scaled to deliver cost-effective foundational sleep training to graduate psychology students. This workshop will accelerate the translation of insomnia management guidelines into psychology practice to improve sleep and mental health outcomes nationwide.

---

Sleep and circadian rhythm disturbances have traditionally been viewed as symptoms or by-products of mental disorders; however, this view is outdated. Sleep and circadian rhythm disturbances share a bidirectional relationship with mental disorders.^1-3^ For example, they predict the onset and course of disorders like depression^4-11^, bipolar disorder^12-16^, post-traumatic stress disorder^17-21^, and psychosis.^2,22-24^ The most common sleep disorder, insomnia, occurs in approximately 50% of people with a mental disorder^25,26^ and has been identified as a robust risk factor for suicide.^27-31^ Conversely, evidenced-based psychological treatments for insomnia, such as Cognitive Behavioral Therapy for Insomnia (CBT-I), effectively improve sleep quality and, in turn, reduce psychopathology.^32-41^ Thus, insomnia is now classified as a disorder in its own right,^42-44^ with clinical practice guidelines worldwide recommending CBT-I as first-line treatment.^45-50^

Despite these updates to practice guidelines, CBT-I has yet to be translated into routine mental healthcare. Mental healthcare providers, such as psychologists, do not view sleep problems as a treatment priority,^51^ nor do they routinely use evidence-based insomnia treatments.^52-54^ This is not surprising, given the ongoing challenges of translating healthcare research findings into clinical practice.^55-57^ Knowledge translation in healthcare (defined as using research evidence to inform health and healthcare decision-making^57^) is slow, with an average of 17 years from publication of high-quality clinical practice guidelines to translation into routine clinical care.^58,59^ While excellent efforts have been made to train mental healthcare providers in sleep and circadian rhythms management,^60-69^ there are major barriers to implementing CBT-I in clinical practice. In particular, the demand for trained CBT-I practitioners far exceeds the supply, with only 752 CBT-I specialists worldwide, predominantly based in the USA.^70-73^ In Australia, only seven out of the 35,315 registered psychologists are internationally recognized as CBT-I specialists,^71,74^ which is insufficient to treat the estimated 3.8 million Australians with insomnia.^75^ Although some digital interventions are starting to be recognized as sufficient for people with uncomplicated insomnia, people with comorbid insomnia and mental health conditions will continue to need interventions provided by mental healthcare providers around the world.^76-81^ Therefore, more psychologists must be trained in CBT-I.

A second barrier to CBT-I dissemination is that our mental healthcare providers lack general sleep and insomnia knowledge due to the limited sleep education taught within university-based healthcare training programs.^52-54,70,82^ Sleep education is scarce in both medical and psychology programs due to limited time, space, and expertise in the curriculum.^69,83-94^ On average, medical students worldwide receive only 2.5 hours of sleep education across their medical degree,^95^ much less than, for example, the 19.6 hours for nutrition education.^96,97^ Sleep education provided to psychology students is even lower. Graduate psychology students in Australia receive a median of only 1 hour of sleep education, with 47% reporting no sleep education at all.^94^ The data are similar in the USA, with only 6% of clinical psychology programs offering a formal course in sleep.^90^ Without foundational sleep education, most graduate psychology students enter the workforce lacking critical CBT-I knowledge and thus incorrectly believing that sleep hygiene is an evidence-based treatment for insomnia.^88,94^

To address these knowledge gaps, sleep education needs to be integrated into the graduate psychology curriculum. Three small studies have developed a sleep education curriculum for graduate psychology programs with promising results.^88,98,99^ For example, we successfully developed and piloted a six-hour behavioral sleep medicine education workshop and observed significant increases in graduate psychology students’ sleep knowledge and self-efficacy to manage insomnia in clinical practice.^99^ Notably, students learned that CBT-I is the first-line treatment for insomnia, not sleep hygiene. Students also provided positive feedback about the workshop, with 100% agreeing that sleep education should be included in all graduate psychology training programs. Sustained clinical behavior change was observed at six months post-workshop, with all students routinely asking their clients about sleep and many using CBT-I components in practice. However, this study was small in scale (*n* = 11) and focused on only one university for a short period (one semester), which is unlikely to bring about sustained change in the number of psychologists with sleep and CBT-I knowledge.

To make large-scale improvements in the translation of CBT-I into mental healthcare, sleep education must be widely integrated into the graduate psychology curriculum. Ideally, this should be aided by a knowledge translation framework to target the barriers to implementation.^67,70,100^ The current study aimed to evaluate the implementation of a large-scale roll-out of our validated behavioral sleep medicine education program, called the Sleep Psychology Workshop^99^, into graduate psychology programs across Victoria, Australia, using the RE-AIM framework.

## Methods

### Study Design

We used a non-randomized waitlist control design, guided by the RE-AIM Framework, to evaluate the implementation of the Sleep Psychology Workshop into all graduate psychology programs in Victoria, Australia in 2020 –2021.

### RE-AIM Framework

The RE-AIM framework (Reach, Effectiveness, Adoption, Implementation, and Maintenance) is widely used in healthcare to evaluate the implementation of evidence-based interventions in the real world and has been used across a variety of settings (e.g., public health, behavioral science, community health, policy).^101,102^ Table 1 outlines the RE-AIM framework dimensions along with how these dimensions were operationalized for the current study.

**Table 1.** A description of the RE-AIM framework dimensions with operationalized definitions.

| RE-AIM Dimension | Definition | Operationalized study definition |
| --- | --- | --- |
| Reach | Reach measures the number, proportion, and characteristics of individuals who are willing to participate in an intervention or program (individual level). | We measured the participation rate, proportion, and characteristics of graduate psychology students willing to attend and participate in the Sleep Psychology Workshop. |
| Effectiveness | Effectiveness investigates the impact of an intervention or program on important individual-level outcomes (e.g., quality of life), including both positive and negative effects (individual level). | For effectiveness, we measured changes in graduate students' sleep psychology knowledge, self-efficacy, preparedness, and confidence to manage sleep disturbances, such as insomnia. |
| Adoption | Adoption refers to the number and proportion of settings who are willing to initiate an intervention (setting level). | We measured the proportion of graduate psychology programs in Victoria, Australia who were willing to adopt the Sleep Psychology Workshop. |
| Implementation | Implementation refers to the consistency and cost of delivering the intervention at the setting level, as well as the clients' use of the intervention and implementation strategies at the individual level (setting and individual level) | For individual level implementation, we explored students' evaluation and feedback about the Sleep Psychology Workshop. At the graduate program level, we reflected on the implementation of the Sleep Psychology Workshop across the different graduate programs and report on the consistency of delivery across sites. |
| Maintenance | Maintenance includes to medium-to long-term sustainability of an intervention or program, such as whether the behavior is sustained by individuals after the intervention and whether the program becomes a part of routine practices or policies at the setting level (setting and individual level). | At the student level, we assessed students' use of the knowledge and skills taught in the Sleep Psychology Workshop 12-months post-workshop. At the program level, we recorded programs interested in continuing the Sleep Psychology Workshop or integrating the content into their curriculum. |

Notes: Table references<sup>101-104</sup>

### Sleep Psychology Workshop: Better Sleep for Better Mental Health Intervention

The Sleep Psychology Workshop: Better Sleep for Better Mental Health is a six-hour interactive online workshop designed to provide graduate psychology students with foundational knowledge and skills in sleep, circadian rhythms, and insomnia management for psychology practice. The six-hour content could be delivered as three, two-hour workshops (normal delivery mode) or two, three-hour workshops (compressed delivery mode) depending on the structure of the graduate psychology program. The development and pilot study of the Sleep Psychology Workshop has been described elsewhere^99^; however in brief, the workshop was developed using a modified Delphi method and contained didactic sleep, circadian rhythms, and CBT-I education, along with interactive/blended learning activities (e.g., completing a sleep diary, sleep restriction therapy calculations), role-play exercises (e.g., taking a sleep history), video (e.g., two-process model of sleep regulation video on YouTube), web-based resources (e.g., Sleep Health Foundation, Sleep Hub), readings from evidence-based literature and textbooks, and handouts with insomnia management exercises (e.g., constructive worry) for clients. The workshop was designed to be foundational in nature, and not train students to CBT-I competency. All lectures were delivered online via Zoom or other university-based video-conferencing software. Interactive activities were conducted via break-out groups between students. A registered psychologist (HM) (master-level qualifications in psychology, internationally certified as a Diplomate in Behavioral Sleep Medicine, six years of clinical experience working as a psychologist in sleep disorder clinics, and 11 years of sleep research experience) delivered the workshop.

### Program selection and participant recruitment

Program coordinators of graduate-level psychology training at Victorian universities in Australia (*n* = 10) were emailed an invitation by the research team to participate in the Sleep Psychology Workshop trial. Program coordinators were advised that the Sleep Psychology Workshop could be run free of charge for their graduate students as part of their coursework requirements or offered as an extra-curricular workshop for students to attend in their own time. Program coordinators were also advised that all graduate students could attend the Sleep Psychology Workshop, but their participation in the research project evaluating the workshop was optional. Ethics committee approvals were obtained for this study (RMIT University CHEAN: 05-19/21939 and Monash University MUHREC: 28828).

Graduate psychology students attending the Sleep Psychology Workshop at their university were invited to participate in the evaluation study. The research team emailed the attending students (details obtained from program coordinators) prior to the workshop to provide information about the voluntary research components of the workshop, along with a copy of the Participant Information and Consent form (also contained within the online study questionnaire). Students were advised that participating in the research study was voluntary and would not impact their university assessments, grades, or relationships with university staff in anyway. Students provided their consent to participate in the evaluation study by ticking a consent box in the online study questionnaire.

## Measures

### Pre-to Post-Workshop

The Graduate Psychology Knowledge, Attitudes, and Practice in Sleep questionnaire (GradPsyKAPS) was used to collect demographic information and measure graduate students’ knowledge, self-efficacy, preparedness and confidence in sleep and insomnia management from pre-to post-workshop (RE-AIM domains = Reach and Effectiveness).^94,99^ A shortened version of the GradPsyKAPS (76 items) was used for the current study to reduce administration time and took approximately 20 minutes to complete (see online supplement). The primary outcome measure for this study was the 35-item multiple choice Sleep Psychology Knowledge Quiz contained within the GradPsyKAPS. The quiz contained items about general sleep and circadian rhythms knowledge (e.g., Current guidelines from the American Academy of Sleep Medicine and Sleep Research Society recommend that per day adults need: (A) at least 6 hours of sleep, (B) at least 7 hours of sleep*, (C) at least 8 hours of sleep, (D) at least 9 hours of sleep) and more clinically-focused insomnia knowledge (e.g., When applying Sleep Restriction Therapy, which is the best response for the Sleep Efficiency (SE) threshold used for increasing prescribed time in bed? (A) 100%, (B) ≥85%*, (C) ≥75%, (D) ≥50%,). The GradPsyKaps was administered through the online platform Qualtrics. To reduce the likelihood of students searching for quiz answers in their notes or online, the quiz was timed, allowing students 45 seconds to answer before progressing to the next question. The order of quiz items was changed post-workshop to reduce learning and order effects.^105^ In addition, the 9-item self-efficacy (7-point Likert scale), the 2-item preparedness (4-point Likert scale) and treatment confidence (4-point Likert scale) subscales from the GradPsyKAPS were used as additional effectiveness outcome measures.

### Post-Workshop

Graduate students in the intervention groups also completed an additional post-workshop survey to provide feedback about the Sleep Psychology Workshop (RE-AIM domain = Implementation). This survey, which took approximately 10 minutes to complete, assessed students’ ratings of the workshop and facilitator, as well as more specific feedback regarding their training experience, learning and workshop content (see online supplement).

### Long-term follow-up

A long-term online feedback survey was administered to students 12 months post-workshop (RE-AIM domain = Maintenance). This measure collected data regarding students’ use of workshop-related sleep psychology knowledge and skills in their clinical practice and took approximately 10 minutes to complete (see online supplement).

### Procedures

Students attended the Sleep Psychology Workshop as part of their graduate coursework or as an extracurricular activity. Students’ participation in the research components of the trial was voluntary. Students were allocated to the intervention or waitlist control group based on the university and semester they attended. Graduate programs running the Sleep Psychology Workshop for the first time were automatically allocated to the intervention group in 2020. Graduate programs who requested repeat administrations of the Sleep Psychology Workshop to different cohorts formed the waitlist control group which ran in 2021. All participants were sent a Participant ID code via email prior to the study to enable the matching of pre-to post-study data and group allocation.

### Intervention Group

The intervention group ran in 2020. All students in the intervention group were emailed information about the research components before they commenced the Sleep Psychology Workshop. If they wished to participate in the research trial, they completed the online pre-workshop questionnaire with consent form via Qualtrics on their mobile device or computer at the beginning of the first workshop (T1). Students who did not wish to participate in the research activities were permitted to do other quiet activities while other students completed the online survey. After completing the Sleep Psychology Workshop, students were asked to complete the online post-workshop questionnaire within two weeks (T2). Twelve months post-workshop, students were invited via email to complete an online follow-up survey.

Students who completed both the pre- and post-workshop GradPsyKAPS questionnaire and scored at least 50% on the Sleep Psychology Knowledge Quiz received a Certificate of Completion for the workshop. In addition, each group of graduate students going through the workshop at their university who completed the pre- and post-assessments went in the draw to win a $50 e-gift voucher for participating in the study. At 12 months post-workshop, all graduate students who completed the long-term follow-up survey could enter a draw to win one of five $100 e-gift vouchers.

### Waitlist Control Group

The waitlist control group ran in the first half of 2021. Graduate students in the waitlist control group were provided with information about the research components via email three weeks prior to the running of the Sleep Psychology Workshop. They were asked to complete the first GradPsyKAPS questionnaire two weeks before the Sleep Psychology Workshop (T1). Students then completed a second administration of the GradPsyKAPS at the beginning of Sleep Psychology Workshop lecture (T2). Students who completed both assessments went in the draw to win a $50 e-gift voucher for participating in the study. If students wished, they could complete the GradPsyKAPS again after the workshop to receive a Certificate of Completion if they scored more than 50% on the Sleep Psychology Knowledge Quiz.

### Data Analysis

Study data were analyzed and visualized using SPSS 28.0 and GraphPad Prism 9.4.1. Descriptive statistics were used to describe the sample. Baseline differences in continuous Sleep Psychology Knowledge Quiz scores were assessed via one-way ANOVA. Changes in pre-to post-workshop sleep psychology knowledge quiz scores were analyzed using mixed between-within ANOVA. For Likert scale ordinal data (e.g., self-efficacy [7-point Likert scale], preparedness [4-point Likert scale], and confidence [4-point Likert scale] subscales) change scores from pre-to post-workshop were calculated and used to compare groups with Kruskal-Wallis tests and post-hoc Dunn’s tests using effect size *r* (*r* = z / √N). Statistical significance level was *p* < .05 (two-tailed). Where specified, a Bonferroni correction was applied to account for multiple post-hoc comparisons between the Waitlist Control, Intervention Groups 1 and Intervention Group 2 (*p* < .017).

Post-workshop evaluation items were reported on a 7-point Likert scale and collapsed into overall agreement, midpoint (neither agree or disagree), and disagreement to simplify data visualization. Descriptive statistics were also used to analyze long-term follow-up data regarding knowledge and skills used by students since completing the workshop. Lastly, students’ free-text items reporting workshop feedback were coded and grouped according to content and frequency of responses (qualitative content analysis).

## Results

### Adoption (Graduate Program Level) and Reach (Graduate Student Level)

Seven out of ten graduate psychology programs across Victoria, Australia agreed to participate in the Sleep Psychology Workshop trial (adoption rate = 70%). In total, the workshop was delivered eleven times across the seven different graduate programs. Four graduate programs included the Sleep Psychology Workshop as part of their graduate coursework requirements, while three offered it as an extracurricular workshop for students to attend independently. Four programs chose the compressed delivery mode (i.e., three by two-hour workshops instead of two by three-hour workshops). For the three non-participating programs, reasons provided for non-participation included: (1) being unable to approve the curriculum change in time for the research project, (2) COVID-19 pandemic challenges, and (3) no response to the study invitation.

In total, 313 graduate psychology students across these seven graduate programs attended the Sleep Psychology Workshop, with 262 consenting to participate in the evaluation study. Eight participants’ data were excluded from the analysis; five due to being enrolled in a Master of Counselling rather that a psychology degree, and three due to missing participant ID codes, leaving 254 for the analysis (Participation rate = 81%). Of students who consented to the trial, 177 completed both pre- and post-workshop assessments (70% study completion rate).

To calculate the true reach of the program to the target population, we compared the number of participants in the intervention group in 2020 (*n* = 172) to the Psychology Board of Australia Registrant data from 2020 to ascertain the number of provisional psychologists enrolled in a Higher Degree Program (*n* = 982) or 5+1 Internship (*n* = 427) in Victoria, Australia (total population *N* = 1409)^106^. The reach of the Sleep Psychology Workshop in 2020 to all provisional psychologists in Victoria studying a Higher Degree Program or 5+1 Internship was 12%.

Group characteristics are displayed in Table 1. Overall, 86% of participants were female, with a mean age of 31 years. Students had received minimal sleep education during their training to date, reporting a median of two hours of sleep education during their undergraduate degree and zero hours in their graduate degree to date. However, 55% of students endorsed already having worked with clients experiencing sleep disturbances whilst on clinical placement or in previous work experience. Most students were completing a Master of Psychology, with 80% in their 5^th^ and 20% in their 6^th^ year of study. As data collection allowed us to capture graduate psychology students at different stages of their training, students completing the Sleep Psychology Workshop were grouped according to their year of study (e.g., Intervention Group 1 = 5th Years, Intervention Group 2 = 6th+ years). Graduate students in the waitlist control group were all in their 5^th^ year of psychology study (Waitlist Group = 5^th^ Years).

**Table 1.** Participant characteristics across the waitlist and intervention groups.

| Variable | Waitlist Control Group<br>(5<sup>th</sup> Year Students)<br>*n* = 82 | Intervention Group 1<br>(5<sup>th</sup> Year Students)<br>*n* = 120 | Intervention Group 2<br>(6<sup>th</sup> + Year Students)<br>*n* = 52 |
| --- | --- | --- | --- |
| **Age (*M* ± *SD*)** | 31.97 ± 9.87 | 30.07 ± 8.08 | 33.11 ± 8.72 |
| **Sex (% female)** | 87 % | 89% | 79% |
| **Year of Study** |  |  |  |
| 5<sup>th</sup> Year | 100% | 100% | 0% |
| 6<sup>th</sup> Year | 0% | 0% | 96% |
| 7<sup>th</sup> + Years | 0% | 0% | 4% |
| **Type of Graduate Program** |  |  |  |
| 5 + 1 Internship* | 41% | 35% | 0% |
| Master of Psychology | 59% | 59% | 83% |
| Doctor of Psychology | 0% | 2% | 11% |
| Combined Masters/PhD | 0% | 4% | 6% |
| **Area of Practice Endorsement** |  |  |  |
| Professional Psychology | 49% | 47% | 0% |
| Clinical Psychology | 0% | 26% | 96% |
| Educational/developmental | 49% | 26% | 0% |
| Community Psychology | 0% | 0% | 4% |
| Health Psychology | 2% | 0% | 0% |
| **Hours of formal didactic sleep**<br>**education [*Med* (range)] during:** |  |  |  |
| Undergraduate degree | 4.00 (0 – 50) | 2.00 (0 – 180) | 2.00 (0 – 16) |
| Honours degree | 0.00 (0 – 50) | 0.00 (0 – 50) | 0.00 (0 – 8) |
| Graduate degree (to date) | 0.00 (0 – 50) | 0.00 (0 – 5) | 1.00 (1 – 20) |

Note: \* the 5 + 1 internship is a new graduate psychology program in Australia and involves completing a one-year accredited Graduate Diploma of Professional Psychology then undertaking a one-year internship of supervised practice before completing the National Psychology Exam. See the Australian Psychology Accreditation Council website. (https://apac.au/students/registration-pathways/) for pathways to registration as a psychologist in Australia.

### Effectiveness (Graduate Student Level)

#### Sleep Psychology Knowledge

Baseline differences in sleep psychology knowledge were assessed via one-way ANOVA and revealed a significant difference between the groups, *F* (2, 219) = 10.33, *p* < .001. Post hoc tests indicated that Intervention Group 2 (6^th^ + years) scored significantly higher on baseline sleep knowledge than both Intervention Group 1 (5^th^ years) (*p* <.001) and Waitlist Control Group (5^th^ Years) (*p* < .001) (see Figure 1). No differences in baseline sleep knowledge were observed between the Waitlist Control Group (5^th^ years) and Intervention Group 1 (5^th^ years) (*p* = .51).

**Figure 1.**
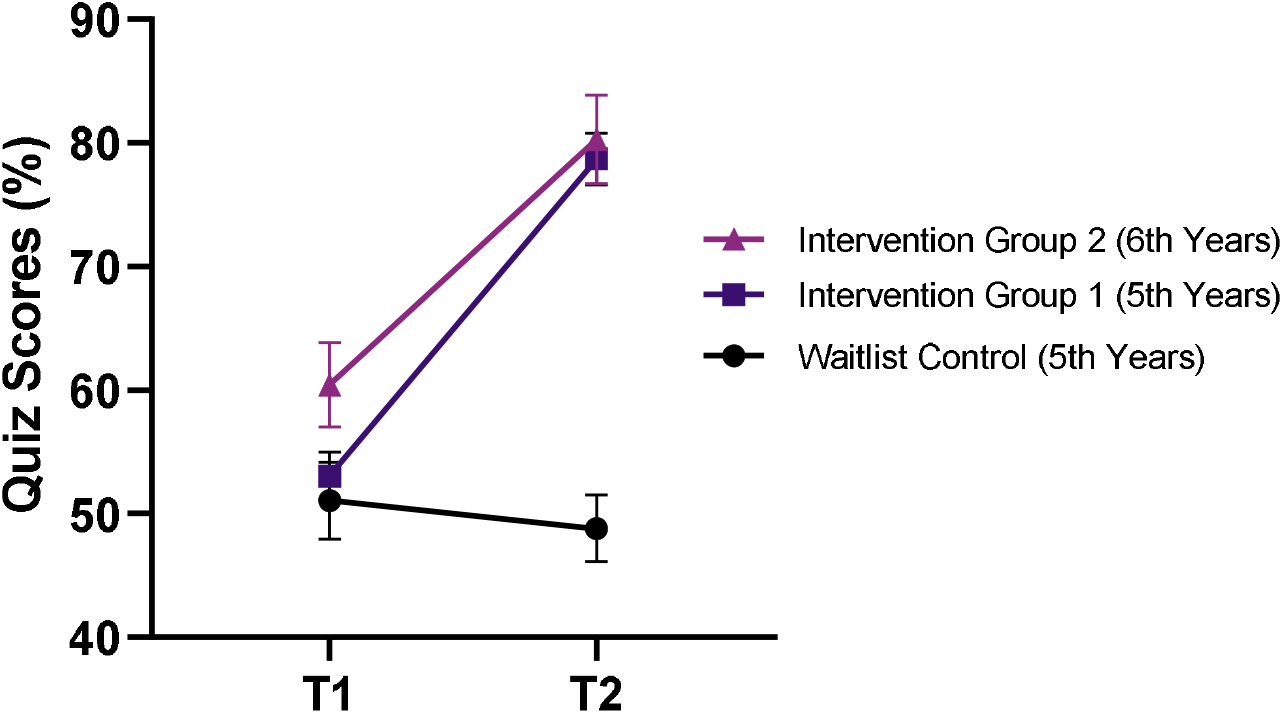
Mean sleep knowledge quiz scores across time for Intervention Group 1 (5th Years), Intervention Group 2 (6th Years) and the Waitlist Control Group (5th Years) (*n* = 175). Error bars indicate 95% confidence intervals.

To assess changes in students’ sleep psychology knowledge from pre-(T1) to post-workshop (T2), a mixed-between within-subjects ANOVA was conducted. There was a significant interaction effect between group and time, Wilks’ Lambda = .47, *F* (2, 172) = 102.78, *p* <.001, partial eta squared = .54, indicating that sleep psychology knowledge quiz results across time differed between the intervention and waitlist groups. There was a significant main effect of time, Wilks Lambda = .37, *F* (1, 172) = 294.03, *p* <.001, with large effect size (partial eta squared = .63), as well as a significant main effect of group, *F* (2, 172) = 57.02, *p* <.001), with large effect size (partial eta squared = .37). Inspection of descriptive statistics and graphs revealed a large increase in sleep psychology knowledge scores for both Intervention Group 1 (5^th^ years) from T1 (*M* = 53.43, *SD* = 10.51) to T2 (*M* = 79.00, *SD* = 10.29) and Intervention Group 2 (6^th^ years) from T1 (*M* = 61.76, *SD* = 10.94) to T2 (*M* = 81.60, *SD* = 9.67). No change was observed in the Waitlist Control Group from T1 (*M* = 51.91, *SD* = 11.68) to T2 (*M* = 50.88, *SD* = 11.08) (see Figure 1).

#### Self-Efficacy, Preparedness, and Confidence in Behavioral Sleep Medicine

Overall, self-efficacy change scores showed significant differences between waitlist control and intervention groups from pre-to post-workshop (all *p* <.001; see Table 2). Both intervention groups reported significantly increased comfort using sleep-related assessment tools and empirically supported treatments, compared to the waitlist group (all *p* <.001). In addition, Intervention Group 1 and 2 reported an increase in their perceived skills to assess and diagnose common sleep and circadian rhythm disorders, compared to the waitlist control (*p* <.001). Similarly, both intervention groups reported increased knowledge of common sleep disturbances seen in mental health disorders, knowing where to go to access further training in sleep, sleep disorders, and circadian rhythms, as well as knowing more about sleep than other graduate psychology students, compared to the waitlist group (all *p* <.01). Both intervention groups reported an increase in how prepared they felt to conduct a sleep evaluation and treat a sleep disturbance using an evidence-based approach (e.g., CBT-I), compared to the waitlist group (*p* <.001, see Figure 2). Lastly, both intervention groups reported significantly increased confidence in treating insomnia disorder and comorbid insomnia using an evidence-based therapy, compared to the waitlist group (*p*’s <.001, see Figure 3).

**Table 2.** Changes behavioral sleep medicine self-efficacy from pre-to post-intervention across groups

|  | Waitlist Control Group |  | Intervention Group 1 |  | Intervention Group 2 |  |  |  |
| --- | --- | --- | --- | --- | --- | --- | --- | --- |
|  | <u>5<sup>th</sup> Years (a)</u> |  | <u>5<sup>th</sup> Years (b)</u> |  | <u>6<sup>th</sup> Years (c)</u> |  |  |  |
| Self-Efficacy Item | Pre-<br>*Median* | Post-<br>*Median* | Pre-<br>*Median* | Post-<br>*Median* | Pre<br>*Median* | Post-<br>*Median* | *p* | Post<br>hoc |
| I feel comfortable using common sleep-related instruments to assess sleep disturbances | 2 | 2 | 2 | 5 | 3 | 6 | <.001 | b, c > a |
| I feel comfortable using empirically supported interventions to treat sleep disturbances | 2 | 2 | 2 | 5 | 3 | 6 | <.001 | b, c > a |
| I have the skills to assess and diagnose common sleep and circadian rhythm disorders | 2 | 2 | 2 | 5 | 3 | 5 | <.001 | b, c > a |
| I know the common sleep disturbances seen in various mental health disorders | 3 | 3 | 3 | 6 | 4 | 5 | <.001 | b, c > a |
| I know where to go to access further training sleep, sleep disorders, and circadian rhythms if required | 2 | 3 | 3 | 6 | 3 | 6 | <.001 | b, c > a |
| I know more about sleep, sleep disorders, and circadian rhythms than most other graduate psychology students | 2 | 2 | 3 | 5 | 3 | 5 | <.001 | b, c > a |

*Note.* The numbers in parentheses in column heads refer to the numbers used for illustrating significant differences in the “Post hoc” test column. Data were collected on a 7-point Likert Scale, from 1 = Strongly Disagree to 7 = Strongly Agree. (*n* = 177) Change scores from post-to pre intervention were used for statistical analysis. A Bonferroni correction was applied to post-hoc Mann-Whitney U Tests with a stricter $\alpha$ level of .017. Effect sizes for post-hoc comparisons between groups b vs a ranged from $r = .56$ to $.73$ , indicating a large effect. Effect sizes for post hoc comparisons between groups c vs a ranged from $r = .39$ to $.60$ , indicating a medium to large effect.

**Figure 3.**
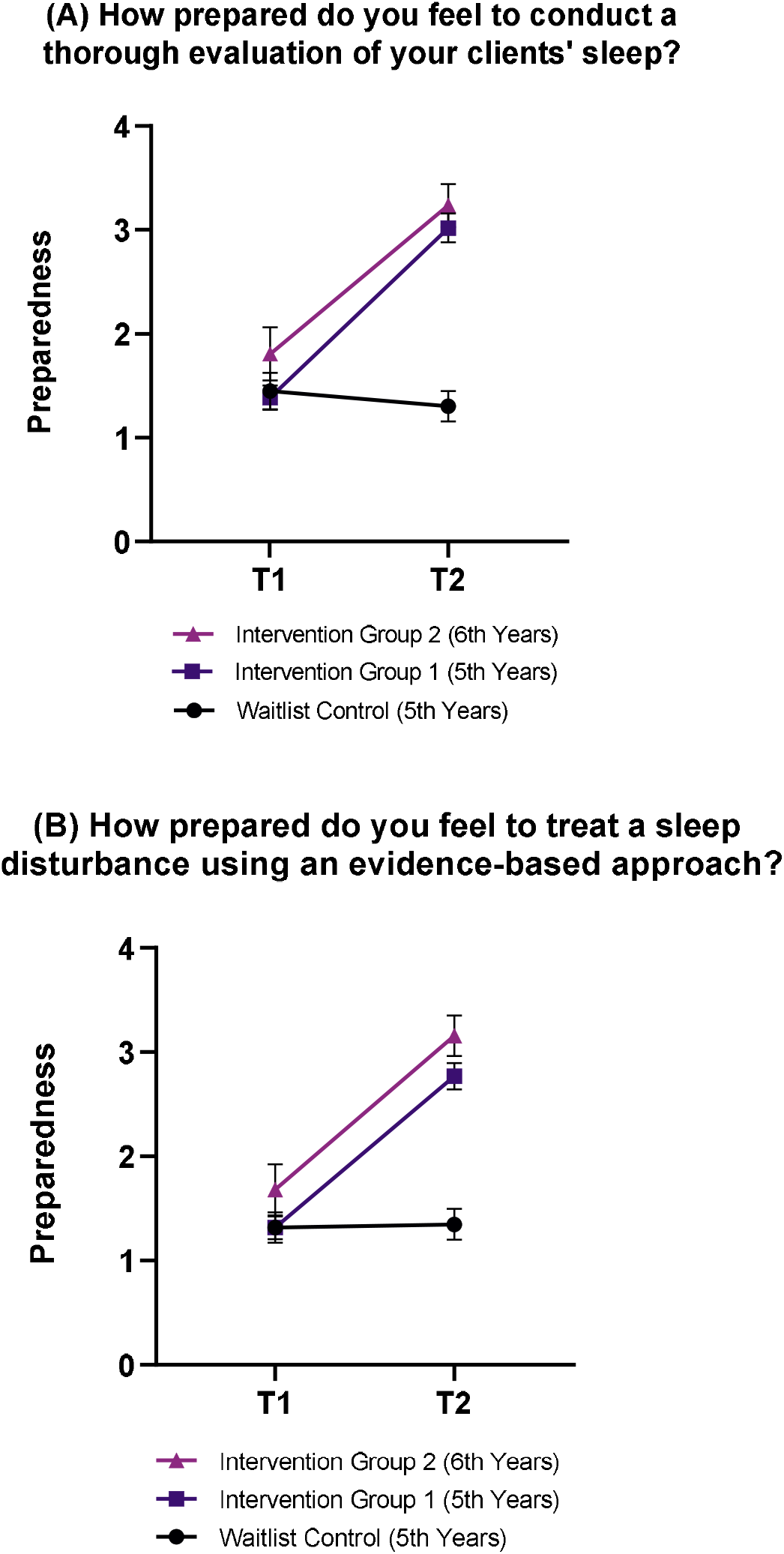
Graduate students’ feelings of preparedness to evaluate (A) and treat (B) sleep disturbances across groups from pre-to post-intervention (*n* = 177). Items were recorded on a 4-point Likert scale from 1 = not prepared to 4 = very prepared. Error bars indicate 95% confidence intervals. A Bonferroni correction was applied to post-hoc Mann-Whitney U Tests with a stricter α level of .017. Effect sizes for post-hoc comparisons between Intervention Group’s 1 and 2 *vs* Waitlist Control ranged from .65 to .83, indicating a large effect.

**Figure 4.**
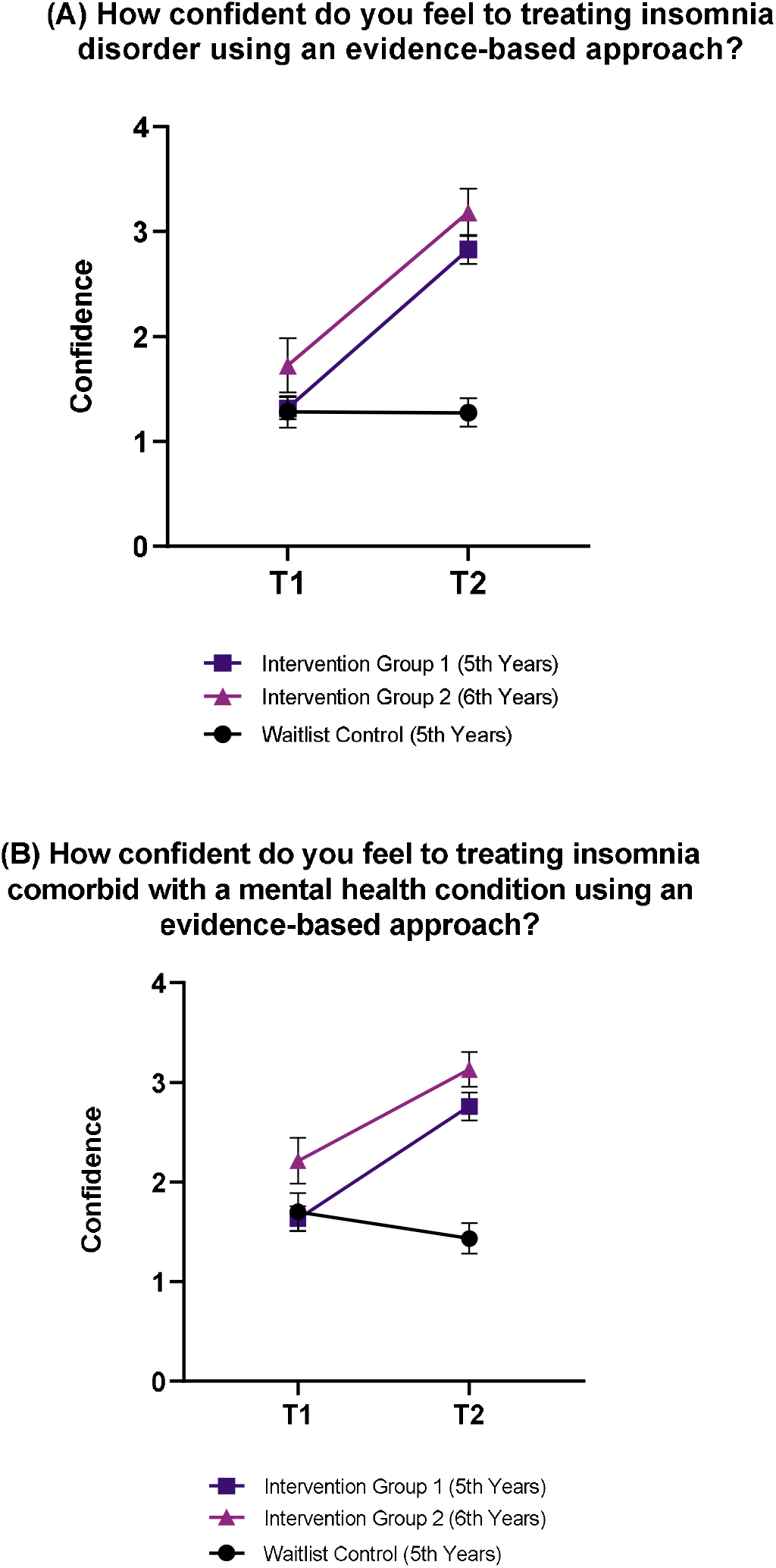
Graduate students’ confidence to treat insomnia disorder (A) and treat comorbid insomnia (B) using an evidence-based therapy (e.g., CBT-I), from pre-to post-workshop (*n* = 177). Items were recorded on a 4-point Likert scale from 1 = not confident to 4 = very confident. Error bars indicate 95% confidence intervals A Bonferroni correction was applied to post-hoc Mann-Whitney U Tests with a stricter α level of .017. Effect sizes for post-hoc comparisons between Intervention Group 1 and2 *vs* Waitlist Control ranged from .59 to .83, indicating a large effect.

### Implementation

#### Graduate Program Level

The Sleep Psychology Workshop was implemented across the different universities using the same study protocol; however, some inconsistencies were noted. Time allocation within the workshop was the main challenge, with some workshop content, mainly the cognitive therapy components of CBT-I, receiving less attention at some universities. For example, more extensive sleep questions by students led to more detailed discussions with some groups than others and consequently reduced the planned didactic content at these sites. In addition, technical issues at one university led to a delay in commencing the workshop, which again rushed some final workshop content. Of note, these time allocation difficulties occurred more at sites which chose the compressed workshop delivery mode (i.e., the universities that chose two, three-hour workshops [*n* = 4] versus three, two-hour workshops [*n* = 3]). However, no differences in students’ post-workshop sleep knowledge quiz scores were identified between the compressed and normal workshop delivery modes (*p* = .098). Lastly, “break out” group work was not possible at one site due to technical difficulties, so role-play exercises were completed as a whole class rather than in pairs.

#### Graduate Student Level

Overall, 138 students from both intervention groups completed the post-workshop evaluation survey (80% completion rate), with 95% of students reporting they attended all 6-hours of the Sleep Psychology Workshop. Students provided positive feedback about the workshop, with 96% rating it as very good-to-excellent (29% = Very Good, 67% = Excellent), and 99% of students also rating the workshop facilitator as very good-to-excellent (11% = Very Good; 88% = Excellent). Additionally, >73% of students endorsed their overall agreement across the different aspects of training experience, learning, and workshop content items (Table 3).

**Table 3.**
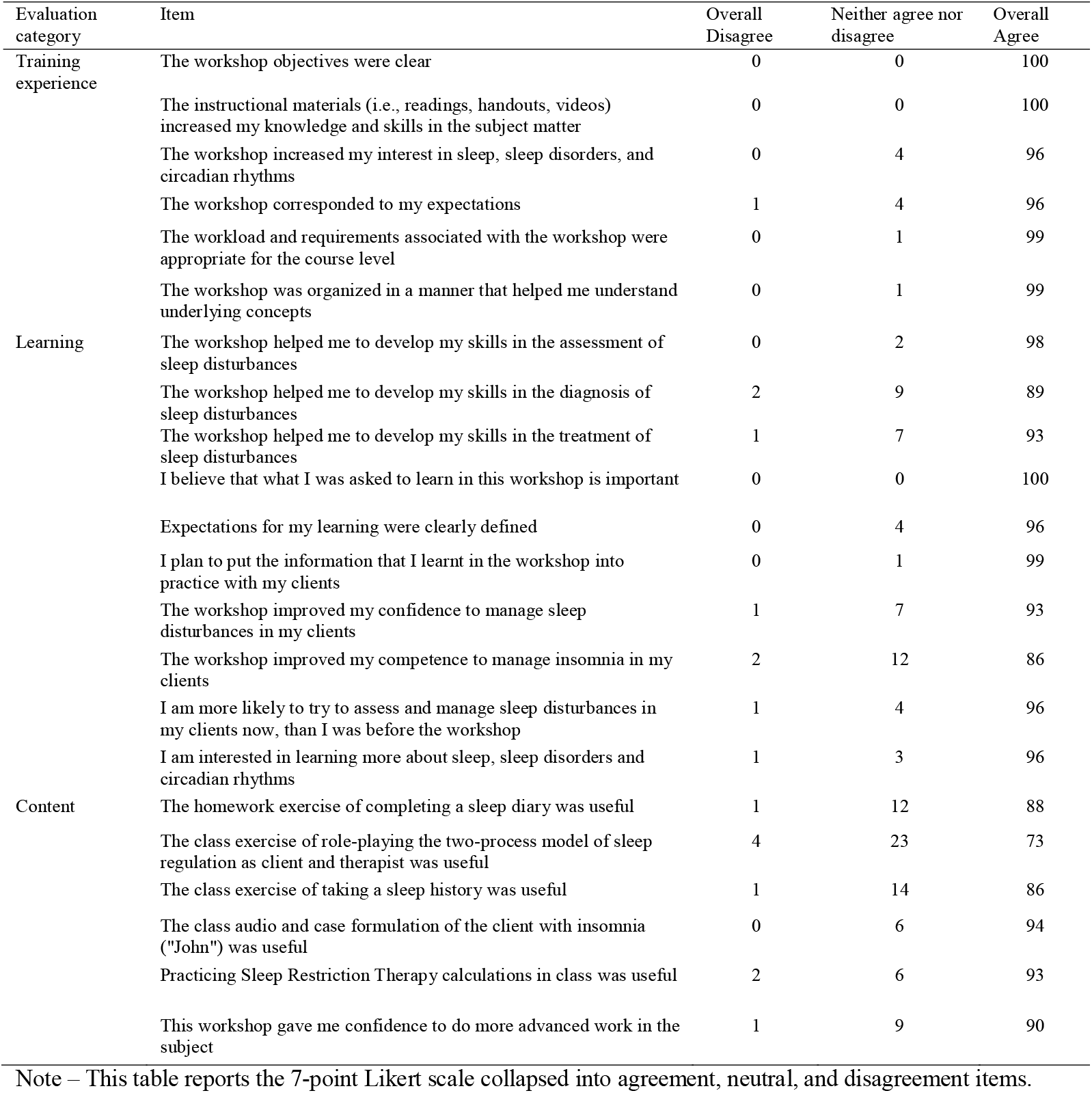
*Evaluation of the Sleep Psychology Workshop (percentage of responses; n = 138)*

Post-workshop, students recognized the importance of sleep, sleep disorders, and circadian rhythms education for psychologists. Overall, 99% of students believed that training in sleep, sleep disorders, and circadian rhythms was important to their future careers as psychologists, with 100% agreeing that all psychologists should understand the relationship between sleep and mental health (see Online Supplement Table S1). Notably, most students endorsed that sleep, sleep disorders, and circadian rhythms (100%), and Cognitive Behavioral Therapy for Insomnia (94%) should be a standard part of the graduate psychology curriculum. Lastly, 96% of students planned to continue learning about sleep, sleep disorders, and circadian rhythms in future, with 75% interested in gaining a professional certification in behavioral sleep medicine once registered as a psychologist and 66% open to studying a postgraduate degree in behavioral sleep medicine.

Twenty-nine students (21% of the post-workshop evaluation sample) provided written feedback on workshop limitations. Nine students commented that the workshops were rushed at the end; seven students commented that they experienced Zoom fatigue with the online workshop format; and six reported they would have preferred three, two-hour workshops, instead of the two, three-hour workshops. Five students also suggested extending the time of the workshop (e.g., to seven hours total). In regard to workshop content, six students requested more interactive activities (e.g., history taking, interventions), more case examples, and time for discussion with the group, with some reduction in the didactic presentation.

### Maintenance of Sleep Psychology Workshop Skills

#### Graduate Program Level

Twelve months after completing the Sleep Psychology Workshop trial, four of the seven graduate programs enquired as to whether the workshop could be delivered to the subsequent years graduate cohort. In addition, two of the graduate programs that did not participate in the research trial contacted the research team requesting the inclusion of the workshop at their university the following year. This indicates an ongoing demand from at least six out of ten universities in Victoria, Australia, for sleep and circadian rhythms education within their programs.

Additionally, one graduate program integrated a shortened version of the Sleep Psychology Workshop into their program coursework (with the researcher’s permission using a “train the trainer” model). This modified version of the Sleep Psychology Workshop formed the basis of a new placement opportunity for students at that university to undertake sleep assessments for members of the community on the University Open Day, and to provide individualized feedback on sleep-wake patterns, sleep duration, chronotype, insomnia symptoms, risk of obstructive sleep apnea, and refer on to appropriate sleep services if required.

#### Graduate Student Level

At 12 months post-workshop, 87 students from the intervention groups responded to a request to complete a long-term follow-up survey about the Sleep Psychology Workshop (Response Rate = 51%). Most students were still finishing their graduate degrees, however, 41% of students reported they had completed their studies. Since completing the Sleep Psychology Workshop, 87% endorsed working with clients experiencing sleep disturbances while on placement/internship or in clinical practice. Insomnia was the most common sleep presentation (71%), followed by nightmares (30%), hypersomnia (31%), circadian rhythms sleep-wake disorders (17%), and obstructive sleep apnea (10%). On average, participants estimated that 46% (SD ± 26%) of their clients reported sleep disturbances and they addressed them with 36% (SD ± 24%) of clients.

Students reported using foundational behavioral sleep medicine skills at the 12-month follow-up, with 100% routinely asking their clients about sleep and 96% feeling comfortable providing psychoeducation about sleep. However, they were less confident in more advanced skills, such as taking a sleep history, referral processes, and delivering evidence-based interventions (see Table 5).

**Table 5.** Self-efficacy in behavioral sleep medicine skills at the long-term follow-up survey

| Item | Overall Disagree | Neither agree nor disagree | Overall Agree |
| --- | --- | --- | --- |
| I routinely ask my clients about sleep | 0 | 0 | 100 |
| I am comfortable providing psychoeducation to clients about sleep | 0 | 4 | 96 |
| I am confident in taking a sleep history with my clients | 8 | 20 | 70 |
| I am aware of when and where to refer clients who require more support with their sleep issues | 17 | 22 | 61 |
| I am comfortable delivering an evidence-based intervention to a client experiencing a sleep disturbance | 13 | 23 | 64 |

Graduate students reported strong implementation and maintenance of the knowledge and skills learned in Sleep Psychology Workshop. Overall, 83% of students endorsed using some of the knowledge and/or skills learned in the workshop on placement or in their clinical practice (see Table 5). However, fewer students had put key behavioral sleep medicine assessment skills into practice, such as using a sleep diary (31%), taking a sleep history (25%), knowledge of common sleep disorders (25%), or administering sleep questionnaires (9%) (see Table 6). For insomnia items, students reported good memory (76%) and clinical application (53%) of the knowledge that spending too long in bed is a common perpetuating factor for insomnia. Students overall reported good memory and application for the components of CBT-I, with 68% using psychoeducation about sleep, 65% using sleep hygiene education, 64% using relaxation training, 47% using stimulus control therapy, and 45% using cognitive therapy in clinical practice. However, only 23% endorsed applying sleep restriction therapy in clinical practice since the completion of the workshop (Table 6).

**Table 6.** Graduate student’s memory and use of sleep knowledge and skills twelve months after completing the Sleep Psychology Workshop.

| Sleep Psychology Workshop Knowledge/skill | I remember this knowledge/skill that was covered in the workshop | I have used this knowledge/skill on placement/internship or in clinical practice |
| --- | --- | --- |
| There is a bidirectional relationship between sleep and mental health problems (e.g., depression) | 79% | 70% |
| Normal sleep physiology (e.g., stages of sleep, normal to wake up at night) | 75% | 45% |
| Sleep recommendations across the lifespan (e.g., at least 7 hours of sleep for adults) | 75% | 58% |
| Consequences of inadequate sleep (e.g., on physical/mental health, cognition) | 79% | 71% |
| The behavioral causes of inadequate sleep (e.g., caffeine, light at night, technology use, irregularity, stress) | 82% | 72% |
| Two-process model of sleep regulation (e.g., Process C and Process S) | 66% | 25% |
| Common sleep disorders (e.g., insomnia disorder, obstructive sleep apnea, circadian rhythm sleep-wake disorders, narcolepsy) | 81% | 25% |
| To ask every client about sleep in your clinical assessment | 77% | 72% |
| Taking a sleep history | 77% | 25% |
| Using a sleep diary | 81% | 31% |
| Common sleep questionnaires (e.g., Insomnia Severity Index, STOP-BANG) | 69% | 9% |
| Insomnia disorder can be an independent or comorbid diagnosis | 66% | 20% |
| Clinical guidelines recommend that you should screen clients who report difficulty initiating and/or maintaining sleep for Insomnia Disorder, even if they have another mental health condition | 43% | 16% |
| Insomnia formulation with the 3P Model (e.g., factors that predispose, precipitate or perpetuate insomnia) | 41% | 12% |
| How and when to refer to a sleep physician | 33% | 9% |
| Spending too long in bed when unable to sleep is a common perpetuating factor of insomnia disorder | 76% | 53% |
| Cognitive Behavioral Therapy for Insomnia (CBT-I) is recommended as first-line treatment for Insomnia Disorder | 69% | 28% |
| The core components of CBT-I are sleep restriction therapy, stimulus control therapy, sleep hygiene education, cognitive therapy, and relaxation training | 70% | 37% |
| Sleep restriction therapy (i.e., improve sleep by reducing time spent in bed to match average total sleep time) | 72% | 23% |
| Stimulus Control Therapy (i.e., bed is for sleep and sex only) | 72% | 47% |
| Sleep Hygiene Education (i.e., maintain a regular sleep-wake routine, don't watch the clock, reduce caffeine) | 76% | 67% |
| Cognitive Therapy (e.g., cognitive restructuring for unhelpful sleep-related thoughts) | 67% | 45% |
| Relaxation training (e.g., progressive muscle relaxation) | 72% | 64% |
| Psychoeducation about sleep and insomnia | 74% | 60% |
| Sleep hygiene education alone is not an effective treatment for insomnia disorder | 61% | 28% |
| Options for further sleep psychology training (e.g., Australasian Sleep Association, Australian Psychological Society Practice Certificate in Sleep Psychology) | 45% | 7% |

#### Final Workshop Feedback

Twelve months on, 91% of students rated the Sleep Psychology Workshop as very good-to-excellent (Very Good = 31%, Excellent = 60%). There was still strong agreement that all graduate psychology students (99%) and registered psychologists (96%) should receive training in sleep, sleep disorders, and circadian rhythms. Notably, 100% of students endorsed that it was important to address sleep disturbances with their clients in clinical practice.

Since workshop completion, 53% reported they had referred to the workshop materials; mainly lecture slides, sleep diaries, and sleep restriction therapy information. In the overall workshop feedback, it was noted that students would have preferred a take-away workshop manual for easy reference. Some students reported completing additional sleep education activities to upskill in CBT-I since completing the workshop, including accessing online sleep information (e.g., Sleep Health Foundation, Sleep Hub; 32%), reading journal articles/textbooks (25%), and individual or peer supervision (22%). Few had taken an online course (5%) or an in-person workshop (1%). However, 38% of students endorsed wanting to receive further training in CBT-I.

## Discussion

This large, controlled trial examined the state-wide implementation of a foundational sleep education workshop for graduate psychology programs using the RE-AIM framework. The Sleep Psychology Workshop had a strong adoption rate by graduate programs (70%), reaching 313 graduate psychology students across the state. Compared to the control condition, the workshop was highly effective at improving graduate students’ sleep psychology knowledge and their self-efficacy, preparedness, and confidence to manage insomnia in clinical practice. Notably, students maintained their sleep knowledge and skills 12 months post-workshop. Students also strongly endorsed that sleep, circadian rhythms, and CBT-I education should be incorporated into all graduate psychology training programs. Overall, this novel sleep education trial measured key implementation outcomes according to the RE-AIM knowledge translation framework^101^ to improve the dissemination of sleep and CBT-I knowledge into mental healthcare.

The strong adoption rate of the Sleep Psychology Workshop demonstrates great interest and demand for sleep education within graduate psychology programs. Not only was the Sleep Psychology Workshop adopted by 70% of programs, after the research trial concluded, 60% of programs reported ongoing demand for the workshop at their university. While most universities wanted the researchers to facilitate ongoing sleep education workshops, one graduate program used a “train the trainer” model to integrate a shortened version of the workshop into their curriculum. Our research demonstrates that graduate programs are willing to adopt a sleep education workshop into their curriculum if facilitated by external experts. Therefore, sustainable annual training models should be established to enable additional sleep education curriculum to be integrated across graduate psychology programs.^100^ Examples could include an educational trainer (funded by professional sleep associations) who delivers the sleep education to university programs, an educational trainer who provides a more in-depth training program for faculty to facilitate the sustainability of the workshop within the institution, or the workshop could be converted into a self-paced digital training program to allow for broader dissemination. Ideally, sleep education should be a hurdle requirement for all graduate psychology students, ensuring that they possess basic behavioral sleep medicine knowledge and skills before graduation.

The Sleep Psychology Workshop reached a meaningful number of future mental healthcare providers (*n* = 313), representing 12% of all graduate psychology students holding a provisional psychology license in Victoria in 2020. However, factors beyond our control limited our ability to capture a higher proportion of graduate students during the study period. Due to time limitations and COVID-19 restrictions, we were only able to run the workshop at some universities for one year level (e.g., 5^th^ years) and so did not reach other enrolled students (e.g., 6^th^ years). Further, students enrolled in the 5 + 1 internship are off campus working full-time for their internship year and may not have been informed of, or able to attend, the Sleep Psychology Workshop. Lastly, Psychology Board of Australia registrant numbers include graduate students who are no longer enrolled in classes (e.g., finishing research components, awaiting degree conferral), limiting their ability to attend the workshop, despite being counted in registrant numbers.^106^ Therefore, future trials of this workshop should ask graduate programs to advertise the Sleep Psychology Workshop to all students, including those who are awaiting degree conferral or out on internship placements to increase reach.

Similar to our pilot study,^99^ and other small sleep education studies,^86,88^ the sleep education intervention was effective at improving graduate students’ sleep and insomnia knowledge, with some educational gains maintained over time. Significant increases in sleep knowledge were found for both 5^th^ and 6^th^ year students, with most students demonstrating retention of basic sleep knowledge 12 months post-workshop. Together, these findings suggest that introducing the Sleep Psychology Workshop early during the 5^th^ year of the graduate school provides trainees with large improvements in important sleep knowledge before commencing clinical placements. Future workshops should also consider additional tangible materials to help students retain and implement sleep knowledge into practice.

Unfortunately, knowledge does not equal competency, nor does knowledge alone lead to behavior change. This was seen in our long-term maintenance data, which identified that not all students had implemented into their clinical practice the sleep-specific skills they had learned. Notably, students reported increased self-efficacy, preparedness, and confidence to manage sleep disturbances, like insomnia, post-workshop. However, while almost all students asked their clients about sleep, only 25% had taken a sleep history and 9% had administered common sleep questionnaires. Further, basic interventions, such as sleep psychoeducation, sleep hygiene, and relaxation training had a higher uptake than more specialized CBT-I skills, such as stimulus control and sleep restriction therapy. These findings are consistent with the training literature, which outlines that didactic training in mental health treatment alone is not always associated with substantial improvements in clinicians’ treatment skills.^107-109^ However, while it is clear that six-hours of sleep education is not enough to achieve CBT-I competency, the Sleep Psychology Workshop is clearly more effective than the minimal sleep education that graduate psychology students currently receive.^94^ More ‘hands-on’ competency training and assessments, integration of sleep interventions on clinical placements, or ongoing professional consultation (e.g., group supervision/telephone consultation) appears crucial for bridging the CBT-I evidence-practice gap.^61,83,107^

Lastly, successful implementation was reflected in the overwhelmingly positive feedback students provided about the quality of the workshop, as well as their learning and training experiences. Further, students were very positive about the importance of sleep and the integration of CBT-I education into all graduate psychology programs. Unlike our pilot study, the role-playing exercise for the two-process model of sleep regulation received a lower quality rating in this study (73% *vs* 87.5% in the pilot study).^99^ This difference is likely a result of differences in implementation, with the pilot study delivered in person (vs. online), allowing the workshop facilitator to listen to students’ role plays and provide prompts/suggestions. It was not possible to ‘drop-in’ on all break-out group role-plays, which limited students’ ability to receive facilitator assistance and feedback, and may have resulted in students not appreciating the full potential of this exercise. In addition, time allocated to different aspects of the workshop (e.g., cognitive therapy for insomnia) differed as a result of time spent engaged in discussions with students over the course of the program. Future workshops should build in additional time for student questions/engagement, adhere more strictly to the timing allotted to each component, and/or increase the time for workshop delivery.

### Strengths

We assessed the real-world implementation of a sleep education workshop within healthcare professional training programs using the validated RE-AIM evaluation framework. By using RE-AIM, we moved beyond individual-level effectiveness data (e.g., changes in students’ sleep knowledge from pre-to post-workshop) to setting-level adoption, implementation, and maintenance data. Setting-level data provided valuable evidence that sleep education can feasibly be implemented into real-world healthcare provider training programs at a state-level. The RE-AIM framework has highlighted the need to focus on the long-term maintenance of the sleep education within graduate psychology programs and provide more practical sleep training opportunities for students to achieve CBT-I competency.

### Limitations

Limitations of this work include that we did not use a randomized-controlled design or include objective clinical competency measures. Future studies would benefit from allocating graduate programs to the intervention or waitlist group in a randomized fashion, ensuring that all groups have an equal distribution of clinical psychology students, as well 5^th^ vs. 6^th^ year students. Additionally, the addition of objective competency assessments would allow for the assessment of students’ practical sleep skills in post-workshop.

### Conclusion and Future Directions

The goal of this innovative study was to widely disseminate an online sleep education workshop to graduate psychology students in Australia. The Sleep Psychology Workshop was effective at both the individual- and setting-level for improving students’ sleep knowledge and skills with a strong adoption rate by graduate psychology programs. To improve the long-term maintenance of this program within graduate psychology programs, we are collaborating with the Australasian Sleep Association (ASA)’s Behavioral Management of Sleep Disorders Education Sub-Committee to strengthen the workshop and drive this dissemination initiative. We also plan to improve the implementation and maintenance of students’ sleep knowledge and skills by offering ongoing group consultation/supervision through a Behavioral Sleep Medicine Online Community of Practice (BSM Online CoP) established through the ASA. Together, we can drive the increase of sleep knowledge and skills within the mental healthcare workforce and build the number of psychologists with CBT-I expertise. For the field of CBT-I dissemination, these are “exciting times, indeed!”^110^

## Data Availability

The data underlying this article will be shared on reasonable request to the corresponding author.

## Acknowledgements

Thank you to the graduate psychology programs and students who participated in the Sleep Psychology Workshop, with special thanks to Andrew Brown and Dianne Summers for their enthusiasm for sleep education for psychologists. We also thank the Australasian Sleep Association (ASA) for endorsing the workshop. Lastly, a huge thank you to the ASA’s Behavioral Management of Sleep Disorders Education Subcommittee, led by Dr Alexander Sweetman, for the upcoming collaboration to disseminate this sleep education workshop widely within Australia! Author 1 is supported by an Australian Government Research Training Program Scholarship administered through Monash University (previously through RMIT University). Monash University’s School of Graduate Research provided funding for participant gift vouchers.

## Data availability statement

The data underlying this article will be shared on reasonable request to the corresponding author.

## Preprint Repositories

