## Supplementary material for "Disseminating sleep education to graduate psychology programs online: A knowledge translation study to improve the management of insomnia": Online Supplement

School of Psychological Science

Monash University

18 Innovation Walk, Clayton, VIC, Australia 3800.

Table S1. Students’ beliefs about behavioral sleep medicine

| Evaluation category | Item | Overall Disagree | Neither agree nor disagree | Overall Agree |
| --- | --- | --- | --- | --- |
| Importance of behavioural sleep medicine | I think training in sleep, sleep disorders and circadian rhythms is important for my future career as a psychologist | 0 | .7 | 99.3 |
|  | I believe all psychologists should understand the relationship between sleep and mental health | 0 | 0 | 100 |
|  | The psychology of sleep, sleep disorders and circadian rhythms should NOT be emphasised in postgraduate psychology training programs | 97.2 | 1.4 | 1.4 |
|  | I believe that learning about behavioral sleep medicine should be a standard part of the postgraduate psychology curriculum | 0 | 0 | 100 |
|  | Cognitive behavioural therapy for insomnia should be a standard part of the postgraduate psychology curriculum | 2.1 | 3.6 | 94.3 |
| Beliefs and intentions for further sleep education | I am interested in receiving further training in the psychology of sleep and sleep disorders | 1.4 | .7 | 97.9 |
|  | I am interested in learning more about sleep, sleep disorders and circadian rhythms | 1.4 | 2.9 | 95.7 |
|  | I plan to continue learning about the psychology of sleep and circadian rhythms | 1.4 | 2.2 | 96.4 |
|  | I would consider studying a postgraduate course in sleep psychology once I am registered as a psychologist | 17.4 | 16.7 | 65.9 |
|  | I am interested in gaining a professional certification in behavioral sleep medicine once I am a registered psychologist | 10.8 | 13.8 | 75.4 |

**Graduate Psychology Students' Knowledge, Attitudes, and Practice in Sleep Questionnaire (GradPsyKAPS – 76 Items)**

**Demographics**

1. Which university are you currently studying your postgraduate psychology degree at?
   a. RMIT University
   b. Latrobe University
   c. The University of Melbourne
   d. Monash University
   e. Victoria University
   f. Federation University
   g. Deakin University
   h. Australia Catholic University
   i. Swinburne University of Technology
   j. ISN Psychology
2. What year of your postgraduate degree are you currently studying? (please complete as full-time student equivalent if you are a part-time student, or just for the master degree component of your degree if you are combined Masters/PhD student)
   a. 5^th^ Year
   b. 6^th^ Year
   c. 7^th^ Year
   d. 8^th^ year
   e. Other (please specify)
3. What type of post-graduate psychology program are you currently enrolled in at your university?
   a. 5 + 1 Internship
   b. Master of Psychology
   c. Doctor of Psychology
   d. Combined Masters/PhD
   e. PhD
   f. Other
4. My training program is preparing me to practice as a registered psychologist or endorsed psychologist with an area of practice endorsement in:
   a. Professional Psychology
   b. Clinical Psychology
   c. Counselling Psychology
   d. Educational and Developmental Psychology
   e. Industrial and Organizational Psychology
   f. Neuropsychology
   g. Community Psychology
   h. Forensic Psychology
   i. Health Psychology
   j. Sport and Exercise Psychology
   k. Other
   l. My degree is primarily research focused and I will not be able to register as a practising psychologist upon completion
5. What is your age?
6. What is your sex?
7. How many hours of formal educational training (i.e., lectures) related to any aspect of sleep, sleep disorders or circadian rhythms have you received so far in your psychology studies approximately? (e.g. one two hour lecture = 2 hours)
   a. Undergraduate degree? : _______
   b. Honours/postgraduate diploma? : _______
   c. Postgraduate degree to date? : _______

**Self-efficacy in sleep, sleep disorders and circadian rhythms.**

Please rate your level of agreement with the following items from (1) strongly disagree to (7) strongly agree.

8. As of today, I feel comfortable using common sleep-related assessment instruments to assess sleep disturbances
9. As of today, I feel comfortable using empirically-supported interventions to treat sleep disturbances
10. I have the skills to assess and diagnose common sleep and circadian rhythm disorders
11. I know the common sleep disturbances seen in various mental health disorders
12. I know where to go to access further training in sleep, sleep disorders and circadian rhythms if required
13. I know more about sleep and circadian rhythms than most other postgraduate psychology students

**Preparedness Scale**

Experience and practice in sleep, sleep disorders and circadian rhythms. Please rate your feelings of preparedness from: (1) Not prepared; (2)	A little prepared; (3) Somewhat prepared; (4) Very prepared.

14. When a client reports symptoms of a sleep disturbance, such as difficulty falling or staying asleep, how prepared do you feel to conduct a thorough evaluation of their sleep?
15. When a client reports symptoms of a sleep disturbance, such as difficulty falling or staying asleep, how prepared do you feel to treat the sleep disturbance using an evidence-based approach?

**Treatment Confidence Scale**

How confident do you feel treating the following sleep problems/disorders using evidence-based therapy? Please rate your feelings of confidence from: (1) Not at all; (2) A little; (3) Somewhat; (4) Very

16. Insomnia disorder
17. Comorbid sleep disturbances in mental health conditions (e.g. sleep disturbances commonly seen in depression)

**Experience working with sleep disturbances.**

18. Have you worked with clients who reported sleep problems during your psychology training or previous work experience? Yes or No

***Intentions for further sleep education***

19. I think training in sleep and chronobiology is important for my future psychology work
20. I am interested in learning more about sleep and chronobiology
21. I believe all psychologists should understand the relationship between sleep and mental health
22. I plan to continue learning about the psychology of sleep and chronobiology
23. The psychology of sleep and chronobiology should NOT be emphasized in postgraduate psychology training programs
24. I am interested in receiving further training in the psychology of sleep and sleep disorders
25. I believe that learning about behavioral sleep medicine should be a standard part of the graduate psychology curriculum
26. Cognitive behavioural therapy for insomnia should be a standard part of the graduate psychology curriculum

**Other comments**

27. Do you have any other comments about your training/education in sleep, sleep disorders or circadian rhythm training for psychology students before you complete the sleep psychology quiz?

**Sleep Psychology Knowledge Quiz**

28. How many people with depression experience sleep and circadian rhythm disturbances?
    a. Up to 30%
    b. Up to 50%
    c. Up to 70%
    d. Up to 90% (correct)
29. It is considered normal to wake up during the night?
    a. Yes (correct)
    b. No
    c. Only in the elderly
    d. Only in young children
44. Which of the following can be considered a sign of insomnia?
    a. Difficulty falling asleep
    b. Difficulty maintaining sleep
    c. Waking up too early
    d. All of the above (correct)
45. Current guidelines from the American Academy of Sleep Medicine and Sleep Research Society recommend that per day adults need:
    a. At least 6 hours of sleep
    b. At least 7 hours of sleep (correct)
    c. At least 8 hours of sleep
    d. At least 9 hours of sleep
46. What is narcolepsy?
    a. A debilitating neurological condition that includes fragmented sleep and daytime sleepiness (correct)
    b. A form of epilepsy impacting on sleep and wake regulation
    c. A factitious disorder where the client believes they cannot stay awake during the day
    d. A result of obstructive sleep apnoea causing excessive daytime sleepiness
47. What is considered a ‘normal’ amount of sleep latency (time to fall asleep)?
    a. No more than 5 minutes
    b. No more than 15 minutes
    c. No more than 30 minutes (correct)
    d. No more than 60 minutes
48. Which of the following is TRUE about obstructive sleep apnoea?
    a. Control of sleep position is the recommended treatment in severe cases
    b. Only men suffer from obstructive sleep apnoea
    c. Continuous positive airway pressure is the gold standard treatment for obstructive sleep apnoea (correct)
    d. Consuming alcohol before bed can improve obstructive sleep apnoea
49. Which of the following is TRUE about obstructive sleep apnoea in children
    a. All children who snore have obstructive sleep apnoea
    b. Children with obstructive sleep apnoea often have behavioural and learning difficulties (correct)
    c. Daytime sleepiness is the primary symptom of obstructive sleep apnoea in children
    d. Most children will grow out of obstructive sleep apnoea so don’t require treatment
50. Initial evaluation of a client with an insomnia complaint should include which of the following?
    a. Polysomnography
    b. Sleep log/Sleep diary (correct)
    c. Beck Depression Inventory
    d. Multiple Sleep Latency Test
51. According to Borbély’s Two-Process Model of Sleep Regulation, homeostatic sleep drive (also known as Process S):
    a. Increases during wakefulness and decreases during sleep (correct)
    b. Increases during a daytime nap
    c. Is regulated by melatonin
    d. Is synchronised by the Earth’s 24 hour rotation cycle
52. Teenagers often go to bed late and, as a result do not get adequate sleep. What is the underlying cause of this delay in teenagers’ bed times?
    a. A delay in their biological clock that occurs during puberty (correct)
    b. An advance in their biological clock that occurs in late adolescence
    c. The need to challenge parental authority through delaying bedtimes
    d. Use of mobile technology in the evening
53. Which of the following most commonly characterises sleep in older individuals?
    a. Sleep need declines significantly
    b. Sleep becomes deeper
    c. The ability to maintain sleep declines (correct)
    d. The frequency of most sleep disorders decreases
54. How long does a person need to go without sleep to produce the same performance deficits as having a Blood Alcohol Content (BAC) of 0.05?
    a. Sleep deprivation does not cause performance impairments like alcohol
    b. 17-19 hours (correct)
    c. 27-29 hours
    d. 37-39 hours
55. Which of the following sleep problem(s) is/are associated with PTSD?
    a. Delayed Sleep-Wake Phase Disorder
    b. Obstructive Sleep Apnoea
    c. Nightmares
    d. All of the above (correct)
56. Which of the following statements about sleep and anxiety is TRUE?
    a. Sleep-related worry only disturbs a persons' sleep if they have an anxiety disorder
    b. Sleep disturbances such as insomnia are not common in anxiety disorders
    c. Trait anxiety is a predisposing factor for insomnia (correct)
    d. Most people with insomnia have an anxiety disorder
57. What are the 5 core components of cognitive behavioural therapy for insomnia?
    a. Sleep hygiene, cognitive restructuring, light therapy, relaxation training, stress management
    b. Sleep restriction therapy, stimulus control, sleep hygiene, cognitive restructuring, relaxation training (correct)
    c. Sleep hygiene, stress management, biofeedback, sleep restriction therapy, behavioural activation
    d. Stimulus control, sleep restriction, light therapy, sleep hygiene, stress management
58. Which of the following is the MOST frequently observed perpetuating or maintaining factor of insomnia disorder (chronic insomnia)?
    a. Poor sleep hygiene
    b. Alcohol use as a sleep aid
    c. Spending too long in bed when unable to sleep (correct)
    d. Not prioritising sleep enough
59. How long does a person need to experience insomnia symptoms with a subsequent impact on daytime functioning to meet the diagnostic criteria for Insomnia Disorder in DSM-5?
    a. One month or more
    b. Three months or more (correct)
    c. Six months or more
    d. Twelve months or more
60. Which treatment for PTSD-associated nightmare disorder is currently the MOST recommended treatment by the American Academy of Sleep Medicine based on the available evidence?
    a. Light therapy
    b. The medication, Prazosin
    c. Imagery rehearsal therapy (correct)
    d. Cognitive Behavioural Therapy
61. According to Mindfulness-Based Therapy for Insomnia, when is the best time to practice mindfulness meditation to improve sleep?
    a. Before bed
    b. At night when you cannot sleep
    c. During the day (correct)
    d. On meditation retreat
62. Screening clients for obstructive sleep apnoea (OSA) is important for psychologists because:
    a. Prevalence studies indicate that people with OSA have higher rates of depression and depressive symptoms than the general adult population
    b. Treatment for OSA can improve mood and cognitive function
    c. Untreated OSA is associated with problems with attention, executive functioning, and memory
    d. All of the above (correct)
63. Common sleep hygiene practices include all of the following EXCEPT...
    a. Refrain from ingesting caffeine within four hours of bedtime
    b. Avoid getting out of bed if you cannot fall asleep (correct)
    c. Avoid watching TV in bed
    d. Avoid taking naps when feeling tired during the day
64. What is sleep efficiency?
    a. How quickly you fall asleep
    b. How rested you feel upon awakening from sleep
    c. The ratio of how much time you sleep relative to how much time you spend in bed (correct)
    d. The ratio of how quickly you fall asleep relative to how much time you spend in bed (correct)
65. If you could only use one strategy with clients to improve insomnia, which of the following has the best evidence as a sole treatment for insomnia?
    a. Sleep hygiene
    b. Biofeedback training
    c. Melatonin
    d. Sleep restriction therapy (correct)
66. Why is the instruction 'Get out of bed if you can't sleep' given to people with insomnia?
    a. To help relax people that cannot sleep
    b. To break the conditioned association of being awake or alert in bed (correct)
    c. To increase motivation to fix the sleep problem
    d. To ensure that people follow strict sleep hygiene instructions
67. Is daytime napping recommended for clients with difficulty initiating sleep?
    a. Yes
    b. No (correct)
    c. Only in the afternoon
    d. Only on days after poor sleep
68. What is one strategy that could assist a person with delayed sleep phase syndrome to re-align their circadian rhythm to an earlier/more desired time?
    a. Recommend they get bright light exposure in the evening
    b. Recommend they get bright light exposure in the morning (correct)
    c. Recommend they set an alarm clock to the desired wake time ASAP
    d. Recommend they see their GP for antidepressant medication
69. According to the principles of stimulus control, what behaviours are permitted in the bedroom?
    a. Sleep and relaxation exercises
    b. Sleep and watching TV
    c. Sleep and reading a paper book
    d. Sleep and sex (correct)
70. What is the best explanation of the rationale for Sleep Restriction Therapy for Insomnia?
    a. In insomnia, there is a mismatch between sleep opportunity and sleep ability. Reducing sleep opportunity will increase homeostatic sleep drive/pressure (correct)
    b. In insomnia, there is a mismatch between sleep opportunity and sleep ability. Increasing sleep opportunity will increase homeostatic sleep drive/ pressure
    c. Sleep restriction therapy restricts a person from sleeping for one whole night to increase homeostatic sleep drive/pressure
    d. People with insomnia are sleep restricted and so sleep restriction therapy gradually increases the time they spend in bed each week to get better sleep.
71. Which is the best response for how to determine the prescribed time in bed for Sleep Restriction Therapy?
    a. How much sleep the client was getting before they had insomnia
    b. The client's average total sleep time plus 30 minutes (correct)
    c. The client's desired total sleep time plus 30 minutes
    d. How much sleep the client reports they need to function well during the day
72. All of the following are evidence-based strategies used in cognitive therapy for insomnia that can help address the perpetuating cognitive elements of insomnia EXCEPT?
    a. Constructive worry
    b. Behavioural experiments
    c. Challenging catastrophic thinking about poor sleep
    d. Counting sheep (correct)
73. When following stimulus control instructions, when is it appropriate to return to the bedroom to go to sleep?
    a. After 15 minutes
    b. When feeling sleepy (correct)
    c. When feeling tired
    d. After eating
74. When applying Sleep Restriction Therapy, which is the best response for the Sleep Efficiency (SE) threshold used for increasing prescribed time in bed?
    a. 100%
    b. ≥85% (correct)
    c. ≥75%
    d. ≥50%
75. What is the best advice for using sleep medication when undergoing Cognitive Behavioural Therapy for insomnia (CBT-I)?
    a. People need to cease all sleep medication prior to commencing CBT-I as it in a contraindication for treatment
    b. Only antidepressant medication can be used as a sleep medication during CBT-I
    c. It is recommended that client's cease sleep medication before the end of CBT-I treatment (correct)
    d. Only melatonin can be used as a sleep medication during CBT-I
76. What is NOT a key element of sleep hygiene?
    a. Ensure the bedroom is dark enough
    b. Limit caffeine intake
    c. Get up at the same time every day
    d. Change your bed sheets regularly (correct)

*Note. The original GradPsyKAPS was published in: Hailey Meaklim, Imogen C. Rehm, Melissa Monfries, Moira Junge, Lisa J. Meltzer & Melinda L. Jackson (2021) Wake up psychology! Postgraduate psychology students need more sleep and insomnia education, Australian Psychologist, 56:6, 485-498, DOI:* [*10.1080/00050067.2021.1955614*](https://doi.org/10.1080/00050067.2021.1955614)*. It was informed by previously published sleep education literature, with permissions obtained from authors to use some items from previous surveys**, including published sleep education and knowledge surveys administered to students in psychology (Peachey & Zelman, 2012), medicine (Almohaya et al., 2013; Mindell et al., 2011; Salas et al., 2013; Sateia, Reed, & Christian Jernstedt, 2005; Stamm, Taylor, Nguyen, & Hardin, 2015; Zozula, Bodow, Yatcilla, Cody, & Rosen, 2001), psychiatry (Khawaja et al., 2017), neurology (Avidan, Vaughn, & Silber, 2013) and pharmacy (Tze-Min Ang, Saini, & Wong, 2008). It also drew upon sleep practice surveys delivered to BSM practitioners (DelGuercio, 2018), psychologists (Zhou, Mazzenga, Gordillo, Meltzer, & Long, 2020) and psychosocial cancer workers (Sweeney & Wu, 2019).*

**Additional survey questions delivered post-workshop**

**Sleep Psychology Workshop Evaluation**
Please indicate your level of agreement with the following statements on a scale from (1) Strongly Disagree to (7) Strongly Agree.

Training Experience

1) The workshop objectives were clear
2) The instructional materials (i.e., readings, handouts, videos) increased my knowledge and skills in the subject matter.
3) The workshop increased my interest in sleep psychology/behavioral sleep medicine
4) The workshop corresponded to my expectations.
5) The workload and requirements associated with the workshop were appropriate for the course level.
6) The workshop was organized in a manner that helped me understand underlying concepts.

Learning

1) The workshop has helped me to develop my skills in the assessment of sleep disturbances.
2) The workshop has helped me to develop my skills in the diagnosis of sleep disturbances.
3) The workshop has helped me to develop my skills in the treatment of sleep disturbances.
4) I believe that what I was asked to learn in this workshop is important
5) Expectations for my learning were clearly defined.
6) I plan to put the information I learnt in the workshop into practice with my clients.
7) The workshop improved my confidence to manage sleep disturbances in my clients.
8) The workshop improved my competence to manage insomnia in my clients.
9) I am more likely to try to assess and manage sleep disturbances in my clients now than I was before the workshop
10) I am interested in learning more about sleep, sleep disorders and circadian rhythms

Content

11) The homework exercise of completing a sleep diary was useful
12) The class exercise of role playing the Two-Process Model of Sleep as client and therapist was useful
13) The class exercise of taking a sleep history was useful
14) The class audio and case formulation of "John" was useful
15) Practicing Sleep Restriction Therapy calculations in class was useful
16) This workshop gave me confidence to do more advanced work in the subject

**Feedback**

17) What overall rating would you give the Sleep Psychology Workshop?
    a. Excellent
    b. Very good
    c. Good
    d. Fair
    e. Poor
18) What overall rating would you give the workshop facilitator?
    a. Excellent
    b. Very good
    c. Good
    d. Fair
    e. Poor

Please indicate your level of agreement with the following statements from (1) strongly disagree to (7) strongly agree.

19) The workshop facilitator demonstrated good knowledge of the subject
20) The facilitator was effective in communicating the content of the workshop
21) The facilitator encouraged feedback from the class
22) The facilitator was enthusiastic about the workshop

Please indicate your level of agreement with the following statements from (1) strongly disagree to (7) strongly agree.

23) I would consider studying a graduate course in sleep psychology/behavioral sleep medicine once I am registered as a psychologist.
24) I am interested in gaining a professional certification in sleep psychology/behavioral sleep medicine one I am a registered psychologist.

**Final questions**

25) Are there any additional resources/materials that would have supported your learning in the Sleep Psychology Workshop or increase your likelihood of applying this learning to your clinical work?
    a. Yes (please specify)
    b. No
26) Was there any area of sleep psychology/behavioral sleep medicine that you would have liked extra training in?
    c. Yes (please specify)
    d. No
27) Do you have any other comments/suggestions for the Sleep Psychology Workshop?

**Long-Term Follow-up Survey Questions**

1) Have you now completed your postgraduate psychology degree?
   a. Yes
   b. No
2) Are you now a registered psychologist with the Australian Health Care Practitioner Regulator (AHPRA)?
   a. Yes - with general registration
   b. Yes - with general registration and currently working towards an area of practice endorsement
   c. Yes - with an area of practice endorsement
   d. Yes - but still with provisional registration
   e. No (please describe)
3) Since completing the Sleep Psychology Workshop, have you worked with any clients on placement/internship or in clinical practice who have experienced sleep disturbances?
   a. Yes
   b. No
   c. Unsure
   d. N/A
4) If yes to (3), what type of sleep disturbances have your observed in your clients? Select all that apply
   a. Insomnia (e.g. difficulties falling/staying asleep)
   b. Disorders of central hypersomnia (e.g., narcolepsy or sleeping long hours and still feeling sleepy/tired
   c. Circadian Rhythm Sleep Wake Disorders (e.g. delayed sleep-wake phase disorder)
   d. Obstructive sleep apnoea (e.g., difficulty breathing when asleep)
   e. Parasomnias (e.g., sleep walking, sleep talking, night terrors)
   f. Nightmares
   g. Restless legs/Periodic limb Movements
   h. Nocturnal panic attacks
   i. Unsure
   j. Other (please describe)
5) Approximately what percentage of your clients experience sleep disturbances? Please provide your best estimate from 0 to 100%
6) Approximately what percentage of your clients do you spend time with addressing sleep disturbances? Please provide your best estimate from 0 to 100%
7) Have you used of the knowledge and/or skills learnt in the Sleep Psychology Workshop on placement or in your clinical practice?
   a. Yes
   b. No
   c. Unsure
   d. N/A

Below is a list of some of the knowledge and skills that were covered in the Sleep Psychology Workshop. Please select if you:

(1) Remember this knowledge/skill that was covered in the Workshop

(2) Have used this knowledge/skill on placement/internship or in clinical practice

Note: Please select all that apply. Please leave the question blank if you do not remember this knowledge/skill and have not used it in practice.

8) There is a bidirectional relationship between sleep and mental health problems (e.g., depression)
9) Normal sleep physiology (e.g., stages of sleep, normal to wake up at night)
10) Sleep recommendations across the lifespan (e.g., at least 7 hours of sleep for adults)
11) Consequences of inadequate sleep (e.g., on physical/mental health, cognition)
12) The behavioral causes of inadequate sleep (e.g., caffeine, light at night, technology use, irregularity, stress)
13) Two-process model of sleep regulation (e.g., Process C and Process S)
14) Common sleep disorders (e.g., insomnia disorder, obstructive sleep apnoea, circadian rhythm sleep-wake disorders, narcolepsy)
15) To ask every client about sleep in your clinical assessment
16) Taking a sleep history
17) Using a sleep diary
18) Common sleep questionnaires (e.g., Insomnia Severity Index, STOP-BANG)
19) Insomnia disorder can be an independent or comorbid diagnosis
20) Clinical guidelines recommend that you should screen clients who report difficulty initiating and/or maintaining sleep for Insomnia Disorder, even if they have another mental health condition
21) Insomnia formulation with the 3P Model (e.g., factors that predispose, precipitate or perpetuate insomnia)
22) How and when to refer to a sleep physician
23) Spending too long in bed when unable to sleep is a common perpetuating factor of insomnia disorder
24) Cognitive Behavioral Therapy for Insomnia (CBT-I) is recommended as first-line treatment for Insomnia Disorder
25) The core components of CBT-I are sleep restriction therapy, stimulus control therapy, sleep hygiene education, cognitive therapy, and relaxation training
26) Sleep restriction therapy (i.e., improve sleep by reducing time spent in bed to match average total sleep time)
27) Stimulus Control Therapy (i.e., bed is for sleep and sex only)
28) Sleep Hygiene Education (i.e., maintain a regular sleep-wake routine, don't watch the clock, reduce caffeine)
29) Cognitive Therapy (e.g., cognitive restructuring for unhelpful sleep-related thoughts)
30) Relaxation training (e.g., progressive muscle relaxation)
31) Psychoeducation about sleep and insomnia
32) Sleep hygiene education alone is not an effective treatment for insomnia disorder
33) Options for further sleep psychology training (e.g., Australasian Sleep Association, Australian Psychological Society Practice Certificate in Sleep Psychology)

Reflecting back on the Sleep Psychology Workshop...

34) What overall rating would you give the workshop?
    a. Excellent
    b. Very Good
    c. Good
    d. Fair
    e. Poor
    f. Unsure/can’t recall
35) What overall rating would you give the workshop facilitator?
    a. Very Good
    b. Good
    c. Fair
    d. Poor
    e. Unsure/can’t recall
36) What was the most helpful thing your learnt by completing the Sleep Psychology Workshop?
37) Is there any aspect of the workshop that could be improved?
38) Have you used or referred back to any of the resources or readings from the Sleep Psychology Workshop?
    a. Yes (please describe which ones)
    b. No
    c. Unsure
39) Are there any additional resources/materials that would have supported your learning in the Sleep Psychology Workshop or increased your likelihood of applying this learning to your clinical work?
    a. Yes (please describe)
    b. No

Please rate your level of agreement/disagreement with the following items from (1) Strongly Agree to (5) Strongly Disagree (select N/A if you have not commenced working with clients on placement yet):

40) I routinely ask my clients about their sleep
41) I am comfortable providing psychology education to clients about sleep
42) I am confident taking a sleep history with my clients
43) I am aware of when and where to refer clients who require more support with their sleep issues
44) I am comfortable delivering and evidence based intervention (e.g., CBT-I) to a client experience a sleep disturbance
45) How important do you think it is to address sleep disturbances with your clients in clinical practice? Please rate from (1) extremely important to (5) not at all important
46) Have you completed any of the following educational activies to upskill in CBT-I since completing the Sleep Psychology Workshop? Please describe
    a. Reading text books/journal articles
    b. Reading online web articles (e.g., Sleep Hub or Sleep Health Foundation websites)
    c. Attended an online course
    d. Attended a workshop
    e. Individual or peer supervision
    f. Other
    g. No but I would like to receive further training
    h. No
47) Is there any other area of sleep, sleep disorders, or circadian rhythms that you would like to receive extra training in?
    a. Yes (please describe)
    b. No

Please rate your level of agreement/disagreement with the following questions from (1) Strongly agree to (5) strongly disagree:

48) All graduate psychology students should receive training in sleep, sleep disorders and circadian rhythms
49) All registered psychologist should receive training in sleep, sleep disorders and circadian rhythms
50) Do you have any other comments about your experience of the Sleep Psychology Workshop?
